# Association between depression and the prevalence and prognosis of prediabetes: a population-based study

**DOI:** 10.1101/2024.05.12.24307252

**Authors:** Jin Zhou, Xiaojiao Yang

**Author notes:** Correspondence: Jin Zhou.

## Abstract

**Background:** Diagnosis and intervention of prediabetes is an emerging approach to preventing the progression and complications of diabetes. It has been reported that inflammatory factors and dysregulation of the hypothalamic-pituitary-adrenal (HPA) axis may be potential pathogenesis mechanisms of diabetes and depression. However, the relationship between depression levels and the prevalence of prediabetes and its prognosis remains elusive. This study aimed to explore the relationship between depression and the prevalence of prediabetes and to further explore the all-cause mortality of different levels of depression in patients with prediabetes.

**Methods:** Our study used a data set from the National Health and Nutrition Examination Survey (NHANES). Participants were divided into two groups (with or without depression) and further divided into subgroups based on different levels of depression status to analyze the relationship between depression and prediabetes prevalence. We then analyzed the relationship between all-cause mortality and depressive status in patients with prediabetes. This study used a weighted multiple logistic/Cox regression model.

**Results:** A total of 4384 participants were included, divided into depression group (n=1379) and non-depression group (n=3005). Results showed that people with depression were at higher risk of developing prediabetes. After adjusting for covariates, moderate to severe depression was positively associated with prediabetes (moderate to severe depression vs no depression: OR=1.834, 95%CI: 0.713–4.721; severe depression vs no depression: OR= 1.004, 95% CI 0.429–2.351). In addition, we explored the relationship between all-cause mortality and depressive status in patients diagnosed with prediabetes (n=2240) and found that moderate to severe depression (HR=2.109, 95%CI 0.952-4.670) was associated with higher mortality in patients with prediabetes. Associated with increased all-cause mortality.

**Conclusions:** Overall, findings suggest that depression is positively associated with prediabetes prevalence and mortality. These results suggest that good management of mental health conditions may be a potential strategy to reduce the occurrence and progression of prediabetes.

## Introduction

Prediabetes is a stage of moderate hyperglycemia with specific parameters including impaired fasting glucose, impaired glucose tolerance (IGT), and specific ranges of hemoglobin A1c (HbA1c) as provided by the American Diabetes Association (ADA). The prevalence of prediabetes has increased over the past few decades, with an incidence rate as high as 43.5% in the United States. Research has shown that people with prediabetes are at higher risk of developing diabetes, cardiovascular complications, and even death. A recent study suggests that early management of prediabetes can reduce the risk of progression to diabetes. Despite this, prediabetes remains under-recognized and under-researched compared to diabetes. Therefore, it is necessary to identify more early intervention measures to prevent the progression of prediabetes to diabetes and the occurrence of its complications. It is well known that traditional complications of diabetes include coronary heart disease, heart failure, stroke, peripheral vascular disease, diabetic nephropathy, retinopathy, and peripheral neuropathy. Although the existence of these complications brings a huge disease burden to patients, as diabetes management continues to improve, the incidence of these complications is gradually declining. Conversely, as patients with diabetes live longer, the risks and burdens of emerging complications of diabetes begin to come into view. ^[1]^ Studies have shown that patients with diabetes have a 25% increased risk of depression. Inflammatory factors and hypothalamic-pituitary-adrenal (HPA) axis disorders may be the underlying pathogenesis of diabetes and depression. ^[2, 3]^ However, whether patients with depression are at increased risk for developing prediabetes and death remains to be determined.

Currently, there is no effective way to avoid the development of prediabetes. Therefore, we tried to find early markers of prediabetes and intervene accordingly. Based on the relationship between mental health conditions and glycemic control, we hypothesized that depression would be associated with a higher prevalence of prediabetes and a worse prognosis. Therefore, the purpose of this study was to determine whether depression is a risk factor for the incidence of prediabetes and adverse outcomes using data from the National Health and Nutrition Examination Survey (NHANES).

## Methods and participants

### Study population

NHANES is an ongoing survey study that provides population estimates related to nutrition and health among adults and children in the United States. The Te survey uses a stratified, multistage probability design to recruit a representative sample of the U.S. population. Data were obtained through personal structured interviews conducted at home, health examinations at mobile examination centers, and sample analysis in laboratories. NHANES 2013-2018 was approved by the National Center for Health Statistics (NCHS) Ethical Review Board and conducted by the 1975 Declaration of Helsinki as revised in 2013. Informed consent was obtained from all participants. Data collection and analysis complied with NHANES research requirements.

### Identification of prediabetes and depressive states

The diagnosis of prediabetes is based on one of the following four principles: (1) Self-reported prediabetes: Participants answered “Have you ever been told about prediabetes/IGT/IFG/borderline diabetes/blood sugar higher than normal, but not high enough? Known as the Diabetes Questionnaire. (2) HbA1c: 5.7%–7.0%. (3) Fasting blood glucose: 5.6-7.0 mmol/L. (4) Oral glucose tolerance test: 7.8-11.0 mmol/L.

The PHQ-9 health questionnaire (Patient Health Questionnaire-9, PHQ-9) is based on 9 items of the diagnostic criteria of DSM-IV (Diagnostic and Statistical Manual of Mental Disorders developed by the American Psychiatric Association). ^[4]^A simple and effective self-rating scale for depressive disorder. It has good reliability and validity in assisting the diagnosis of depression and assessing the severity of symptoms. Participants answered this questionnaire based on the situation in the past two weeks. The answers to each question From left to right are “rarely”, “a few days”, “more than half”, and “almost every day”. The scores of the corresponding options are: 0, 1, 2, 3. Add each question to get the final score.

### Inclusion and exclusion process

We used data from the NHANES 2013-2018 cycle. First, we included 4384 patients with clear data on depression status, prediabetes, and other covariates to explore the relationship between depression status and risk of prediabetes. In addition, we also recruited 2240 prediabetic patients from a sample of 4384 to further explore the relationship between depressive status and all-cause mortality (Fig. 1).

**Fig. 1.**
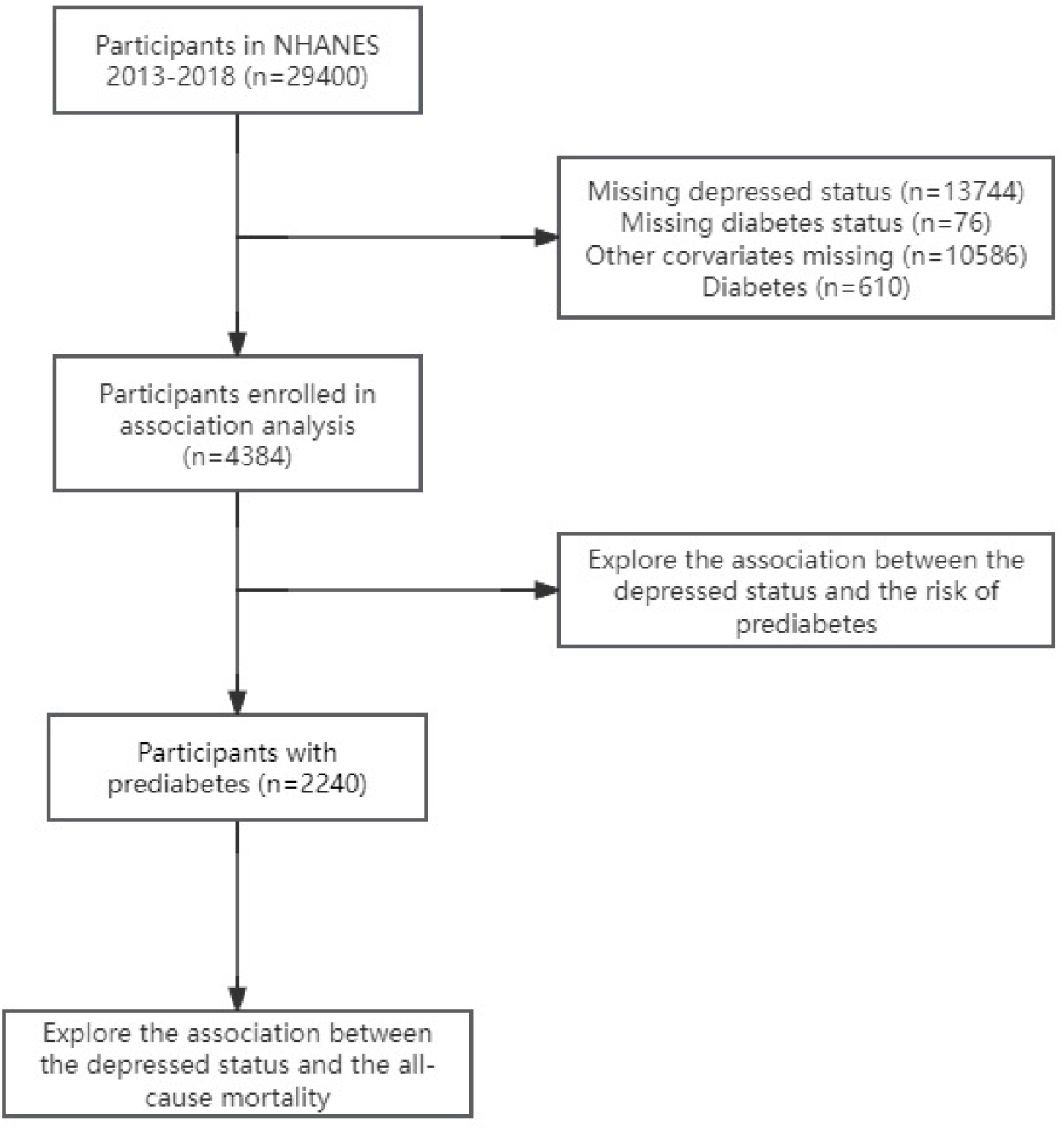
Study workflow

### Outcomes

The result of the correlation analysis was a diagnosis of prediabetes. The outcome of the survival analysis was mortality among patients diagnosed with depression in prediabetes. All-cause mortality was determined through linkage to National Death Index records for 2019. ICD-10 is used to determine cause of death.

### Assessment of covariates

A standardized questionnaire obtained information on sociodemographic characteristics, smoking status, and alcohol consumption. Individuals who had smoked >100 cigarettes in their lifetime were considered current smokers, whereas those who had smoked >100 cigarettes and had quit smoking were considered former smokers. A never-drinker does not drink alcohol every year, a light drinker drinks 1-11 times a year, a moderate drinker drinks 1-3 times a month, and a moderate drinker drinks 1-4 times a week. Heavy drinkers are those who drink alcohol almost every day. Body mass index (BMI) is calculated from weight/height[2] (kg/m[2]). The diagnosis of diabetes is as follows: (1) The doctor diagnoses you as having diabetes; (2) hemoglobin A1c>7%; (3) Fasting blood glucose ≥7.0 mmol/L; (4) Random blood glucose ≥11.1 mmol/L, (5) Oral administration within 2 hours Glucose tolerance test (OGTT) blood glucose ≥11.1 mmol/L; (5) Use of diabetes drugs or insulin. Hypertension, hyperlipidemia, and coronary heart disease (CHD) were identified as relevant diseases with “yes” responses to the questionnaire. In addition, the researchers conducted relevant laboratory data analysis, including assessment of baseline glucose, insulin, hemoglobin A1c, total cholesterol, triglycerides, high-density lipoprotein, and low-density lipoprotein levels.

### Statistical Analysis

All analyses incorporate sample weights, strata, and primary sampling units to produce accurate national estimates. Sample characteristics are reported as means (SEs) for normally distributed continuous variables, medians for non-normally distributed continuous variables, and percentages for categorical variables. Using the no-depression group as the reference, a weighted multivariable logistic regression model was used to estimate the OR and 95% confidence interval associated with depressive status and prediabetes. Weighted Cox proportional hazards regression was used to estimate hazard ratios (HRs) and 95% CIs for all-cause mortality associated with depressive status. Person-time was calculated as the time interval between the NHANES interview date and the date of death or end of follow-up (December 31, 2019), whichever occurred more frequently. We constructed three statistical models. Model 1: No adjustment for covariates. Model 2: For age (continuous), sex (male or female), race/ethnicity (Mexican American, other Hispanic, non-Hispanic white, non-Hispanic black, or other race), and education level (less than high school, High school, high school and above) have been adjusted. Model 3: Age (continuous), sex (male or female), race/ethnicity (Mexican American, other Hispanic, non-Hispanic white, non-Hispanic black, or other race), education level (less than high school, high school, high school or above), smoking status (former or current), alcohol consumption (never, light, moderate, moderate to severe, severe), BMI (continuous), hypertension (yes or no), hyperlipidemia (yes or no) ) and coronary heart disease (yes or no) were adjusted. Individuals with missing covariate data in Model 2 were not included in the analyses of missingness in Model 2 and Model 3 (listwise deletion). Additionally, stratified analyses were performed to examine whether the detected associations differed by age, sex, and body mass index. All analyses were performed using R software (4.1.0). A two-sided *p*<0.05 was considered statistically significant.

## Results

### Participant Characteristics

In this study, 4384 participants participated in our analyses. The patients were 1379 patients with depression and 3005 patients without depression. Compared with participants without depression, participants with depression were more likely to be female, have lower education, have hypertension, hyperlipidemia, moderate to heavy alcohol use, current smoking, higher BMI, and have diabetes Early stage.

### Logistic regression between depression and prediabetes

We used a weighted multivariable logistic regression model to explore the relationship between depression and prediabetes (Table 2). Compared with non-depression, depression was positively associated with prediabetes in all models (OR=1.023, 95% CI 0.901-1.162; 1.117, 95% CI 0.976-1.279; 1.016, 95% CI 0.881-1.172). All models have *p*-values less than 0.001. According to the PHQ-9 Health Questionnaire, we further divided depressive states into 5 types. In model 1, all depressive states were significantly associated with prediabetes (OR=1.023, 95% CI 0.901–1.162; 1.117, 95% CI 0.976–1.279; 1.016, 95% CI 0.881–1.172). The *p*-values for all depression states were less than 0.001. After adjusting for covariates, in model 2 (OR=1.065, 95% CI 0.735–1.544; 2.076, 95% CI 0.831–5.185) and model 3 (OR=1.029, 95% CI 0.594–1.782; 1.834, 95% CI 0.713 –4.721; 1.004, 95%CI 0.429-2.351), moderate to severe and severe depression were positively associated with prediabetes. Furthermore, significant associations remained between depression and prediabetes in most subgroups.

**Table 1.**
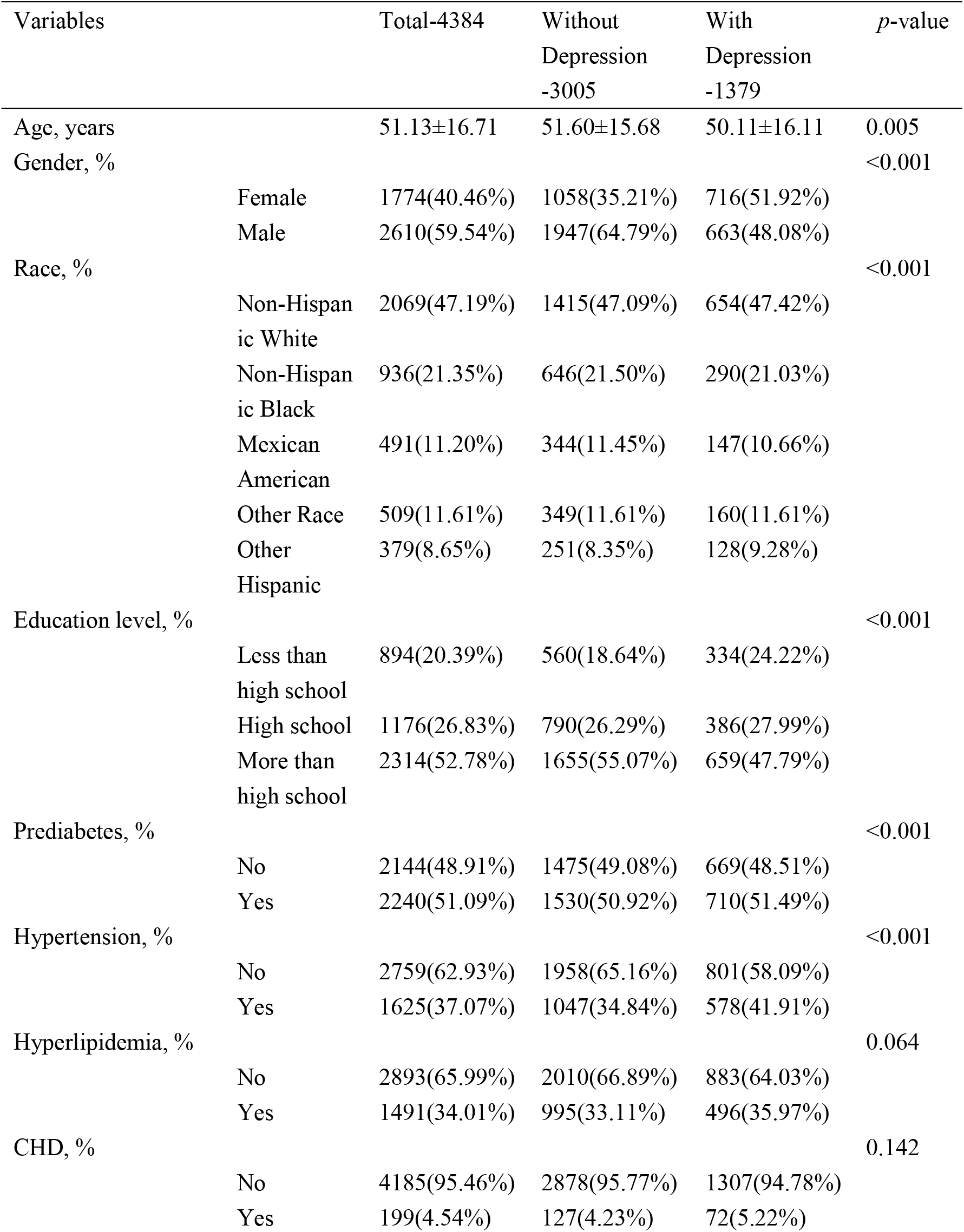
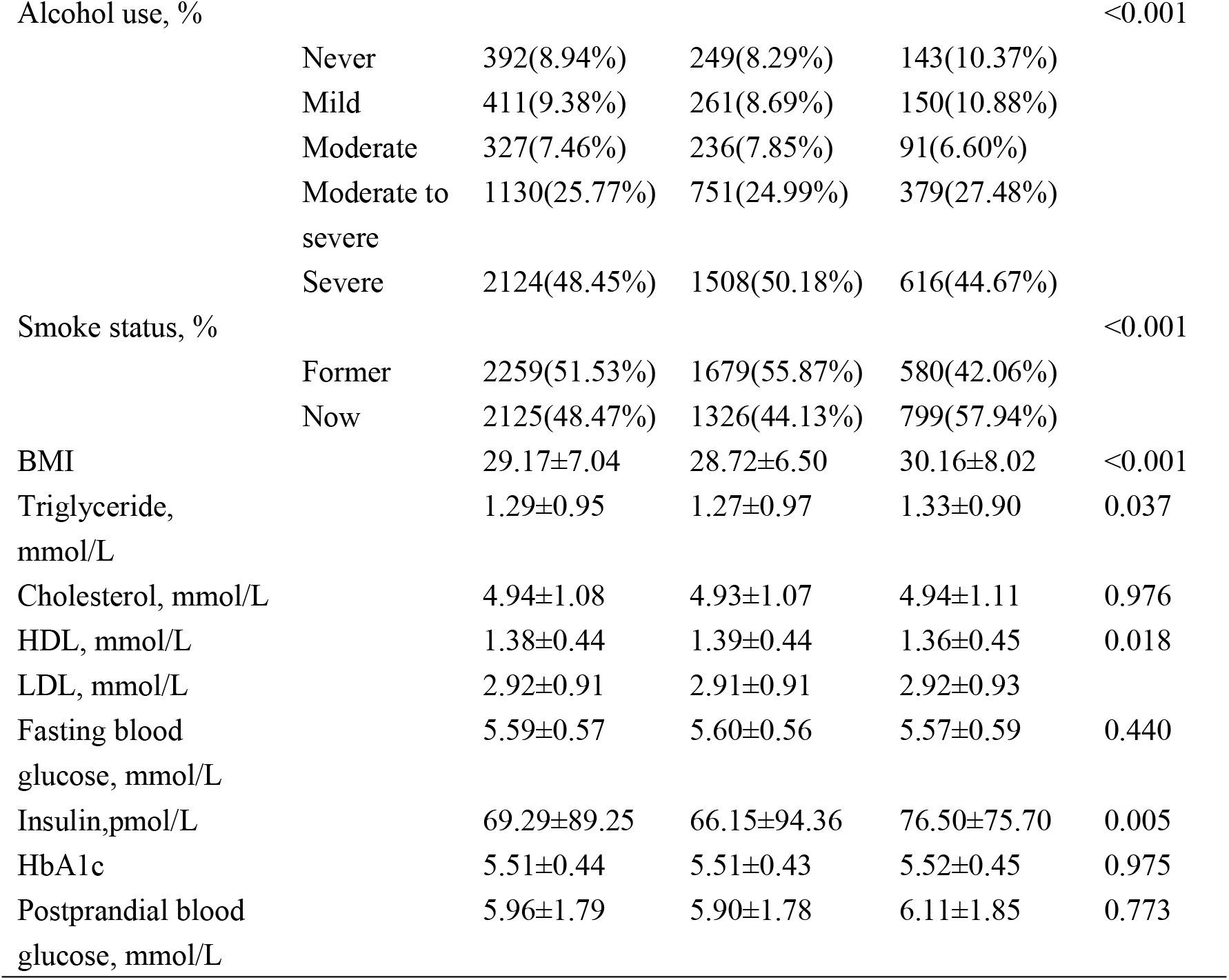
Baseline characteristics of included participants in NHANES 2013-2018.

**Table 2.**
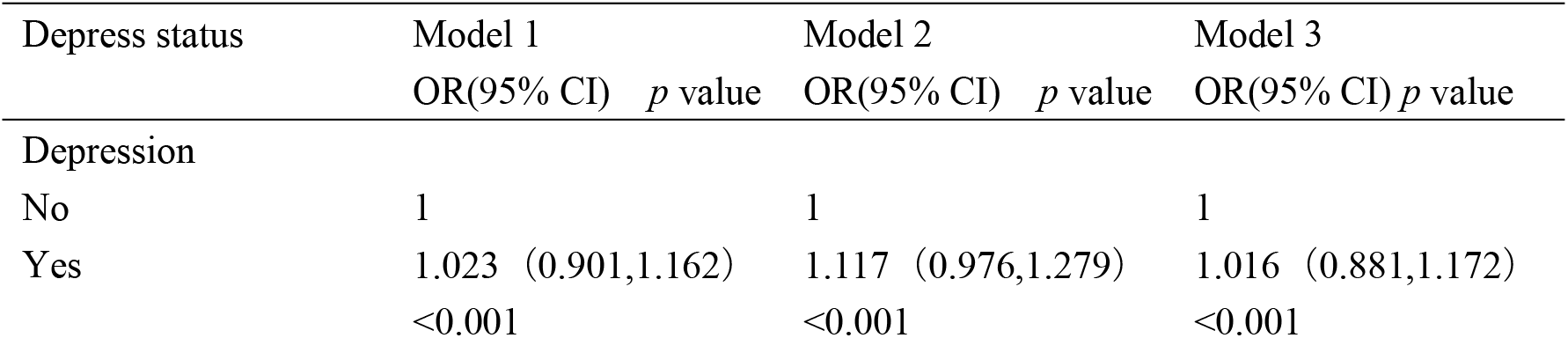
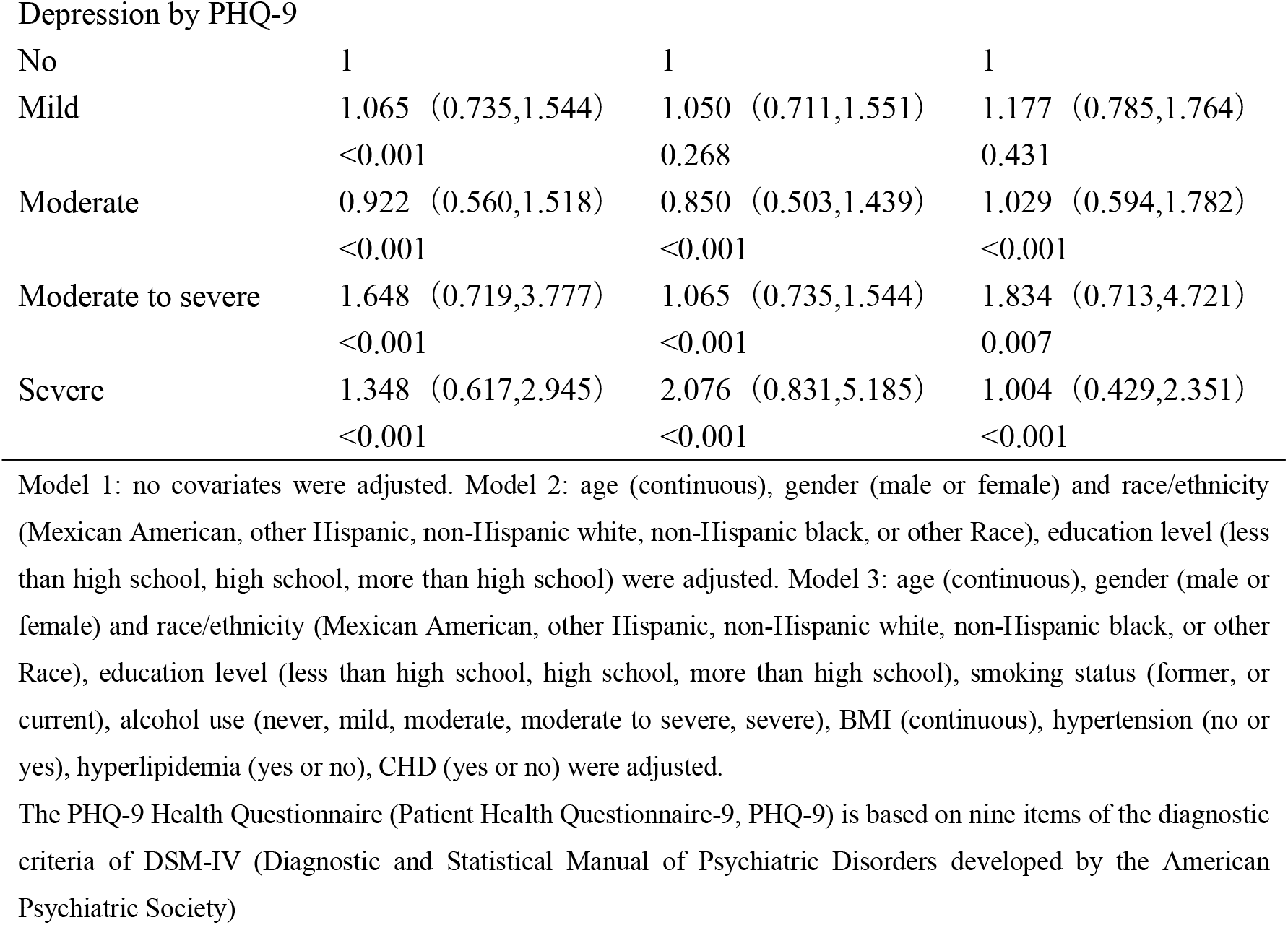
Association between prediabetes and depression among participants in NHANES 2013-2018.

| Depress status | Model 1 | Model 2 | Model 3 |
| --- | --- | --- | --- |
|  | OR(95% CI) *p* value | OR(95% CI) *p* value | OR(95% CI) *p* value |
| Depression |  |  |  |
| No | 1 | 1 | 1 |
| Yes | 1.023 (0.901,1.162)<br><0.001 | 1.117 (0.976,1.279)<br><0.001 | 1.016 (0.881,1.172)<br><0.001 |
| Depression by PHQ-9 |  |  |  |
| No | 1 | 1 | 1 |
| Mild | 1.065 (0.735,1.544)<br><0.001 | 1.050 (0.711,1.551)<br>0.268 | 1.177 (0.785,1.764)<br>0.431 |
| Moderate | 0.922 (0.560,1.518)<br><0.001 | 0.850 (0.503,1.439)<br><0.001 | 1.029 (0.594,1.782)<br><0.001 |
| Moderate to severe | 1.648 (0.719,3.777)<br><0.001 | 1.065 (0.735,1.544)<br><0.001 | 1.834 (0.713,4.721)<br>0.007 |
| Severe | 1.348 (0.617,2.945)<br><0.001 | 2.076 (0.831,5.185)<br><0.001 | 1.004 (0.429,2.351)<br><0.001 |

Model 1: no covariates were adjusted. Model 2: age (continuous), gender (male or female) and race/ethnicity (Mexican American, other Hispanic, non-Hispanic white, non-Hispanic black, or other Race), education level (less than high school, high school, more than high school) were adjusted. Model 3: age (continuous), gender (male or female) and race/ethnicity (Mexican American, other Hispanic, non-Hispanic white, non-Hispanic black, or other Race), education level (less than high school, high school, more than high school), smoking status (former, or current), alcohol use (never, mild, moderate, moderate to severe, severe), BMI (continuous), hypertension (no or yes), hyperlipidemia (yes or no), CHD (yes or no) were adjusted.
The PHQ-9 Health Questionnaire (Patient Health Questionnaire-9, PHQ-9) is based on nine items of the diagnostic criteria of DSM-IV (Diagnostic and Statistical Manual of Psychiatric Disorders developed by the American Psychiatric Society)

### Cox regression analysis between depression and mortality in patients with prediabetes

A total of 2240 patients with prediabetes were included, and the weighted Cox proportional hazards regression method was used to further explore the correlation between depression status and mortality in patients with prediabetes. The average follow-up time was 44.47±43.84 months. Compared with the non-depressed group, patients with depressed status were significantly associated with mortality in prediabetes in all models (HR=1.570, 95% Cl 1.180–2.088, *p*=0.002; 1.816, 95% Cl 1.359–2.427, *p* <0.001; 1.708, 95% Cl 1.273–2.290, *p*<0.001). After stratifying by depression status, moderate and moderate-severe depression were positively associated with prediabetes mortality (HR=1.946, 95% Cl 1.069–3.542, *p*=0.029; 2.682, 95% Cl 1.283–5.605, *p*=0.009). After further adjusting for age, gender, race, education level, drinking status, smoking status, BMI, hypertension, hyperlipidemia, and coronary heart disease, moderate to severe depression was associated with model 2 (HR=2.459, 95% CI 1.118– 5.407, *p*=0.025) and model 3 (HR=2.109, 95%CI 0.952–4.670, *p*=0.032) were related to prediabetes mortality (Table 3), and the Kaplan-Meier survival curve also obtained similar results (Figure 2).

**Table 3.**
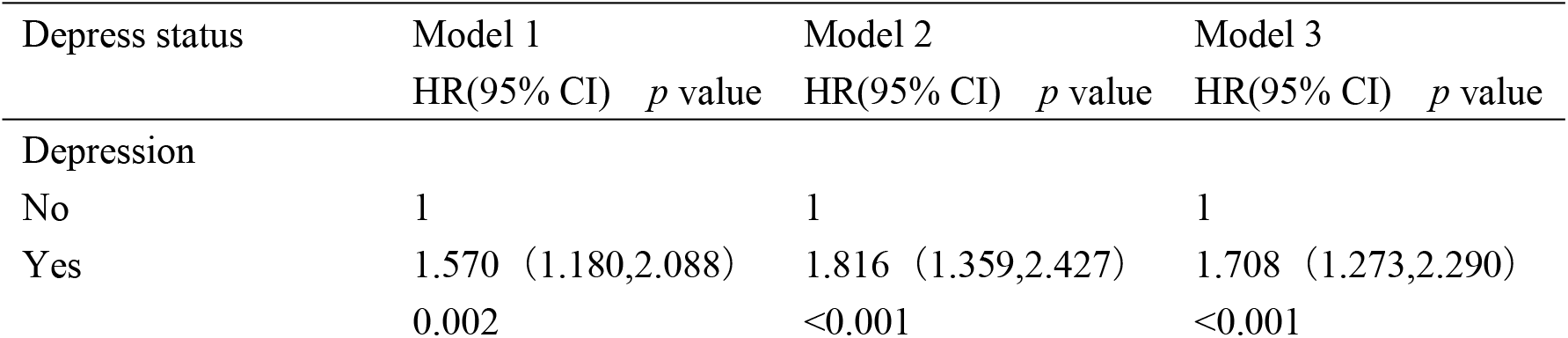
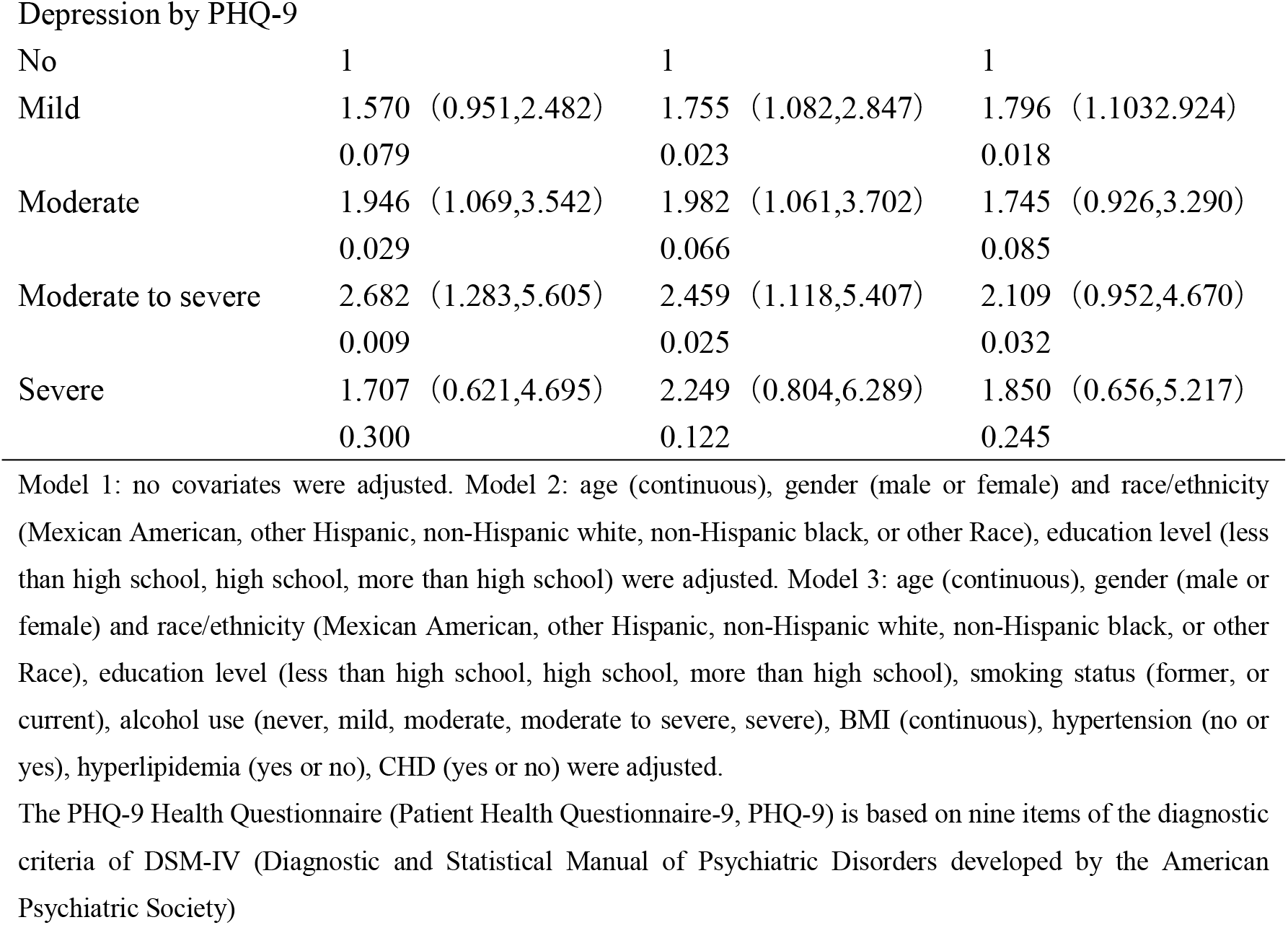
All-cause mortality of patients with prediabetes during different depress status in NHANES 2013-2018.

| Depress status | Model 1 |  | Model 2 |  | Model 3 |  |
| --- | --- | --- | --- | --- | --- | --- |
| | HR(95% CI) | $p$ value | HR(95% CI) | $p$ value | HR(95% CI) | $p$ value |
| Depression |  |  |  |  |  |  |
| No | 1 |  | 1 |  | 1 |  |
| Yes | 1.570 (1.180,2.088)<br>0.002 |  | 1.816 (1.359,2.427)<br><0.001 |  | 1.708 (1.273,2.290)<br><0.001 |  |

Depression by PHQ-9
|  |  |  |  |
| --- | --- | --- | --- |
| No | 1 | 1 | 1 |
| Mild | 1.570 (0.951,2.482)<br>0.079 | 1.755 (1.082,2.847)<br>0.023 | 1.796 (1.103,2.924)<br>0.018 |
| Moderate | 1.946 (1.069,3.542)<br>0.029 | 1.982 (1.061,3.702)<br>0.066 | 1.745 (0.926,3.290)<br>0.085 |
| Moderate to severe | 2.682 (1.283,5.605)<br>0.009 | 2.459 (1.118,5.407)<br>0.025 | 2.109 (0.952,4.670)<br>0.032 |
| Severe | 1.707 (0.621,4.695)<br>0.300 | 2.249 (0.804,6.289)<br>0.122 | 1.850 (0.656,5.217)<br>0.245 |

Model 1: no covariates were adjusted. Model 2: age (continuous), gender (male or female) and race/ethnicity (Mexican American, other Hispanic, non-Hispanic white, non-Hispanic black, or other Race), education level (less than high school, high school, more than high school) were adjusted. Model 3: age (continuous), gender (male or female) and race/ethnicity (Mexican American, other Hispanic, non-Hispanic white, non-Hispanic black, or other Race), education level (less than high school, high school, more than high school), smoking status (former, or current), alcohol use (never, mild, moderate, moderate to severe, severe), BMI (continuous), hypertension (no or yes), hyperlipidemia (yes or no), CHD (yes or no) were adjusted.
The PHQ-9 Health Questionnaire (Patient Health Questionnaire-9, PHQ-9) is based on nine items of the diagnostic criteria of DSM-IV (Diagnostic and Statistical Manual of Psychiatric Disorders developed by the American Psychiatric Society)

**Fig. 2.**
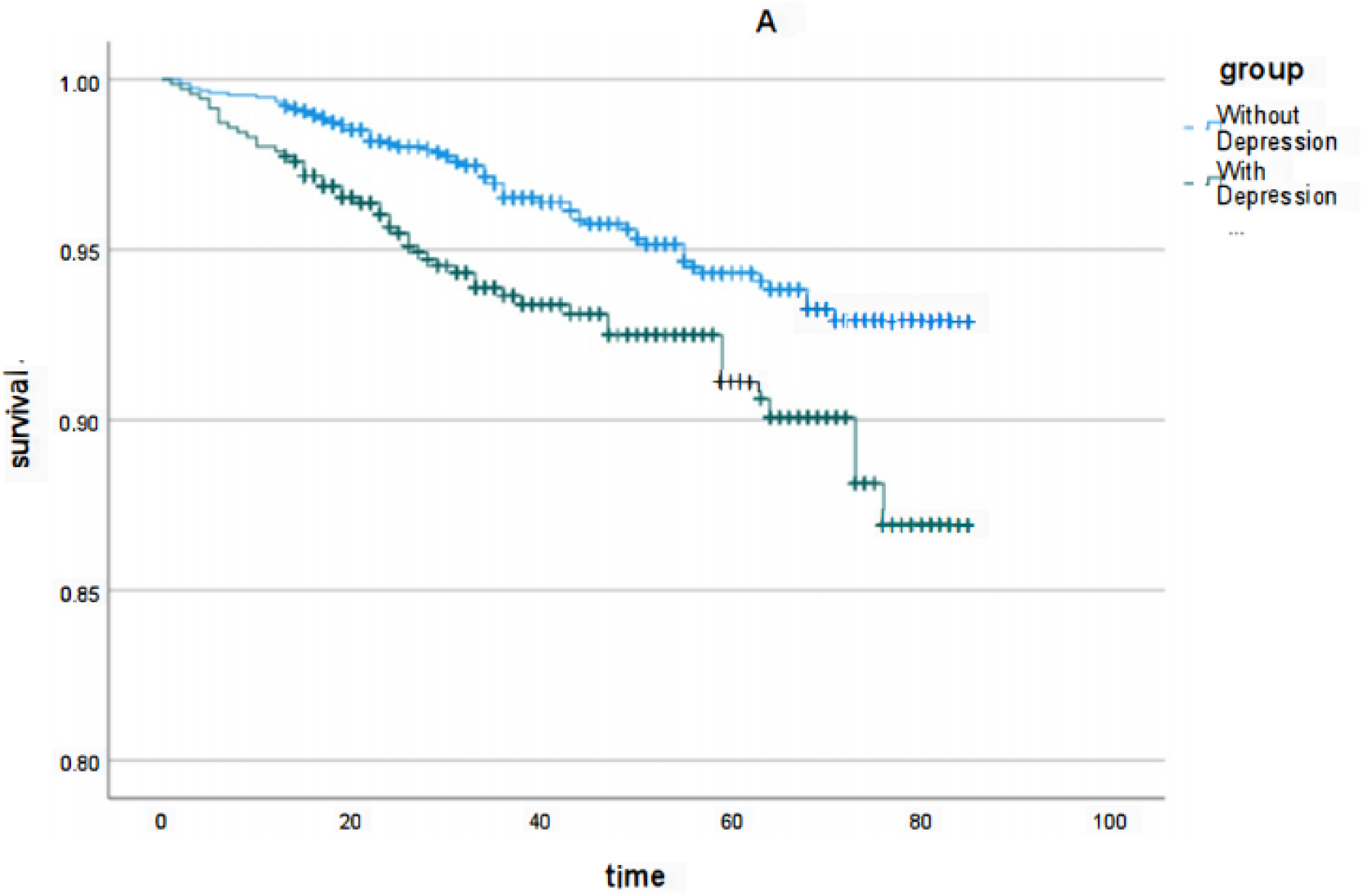
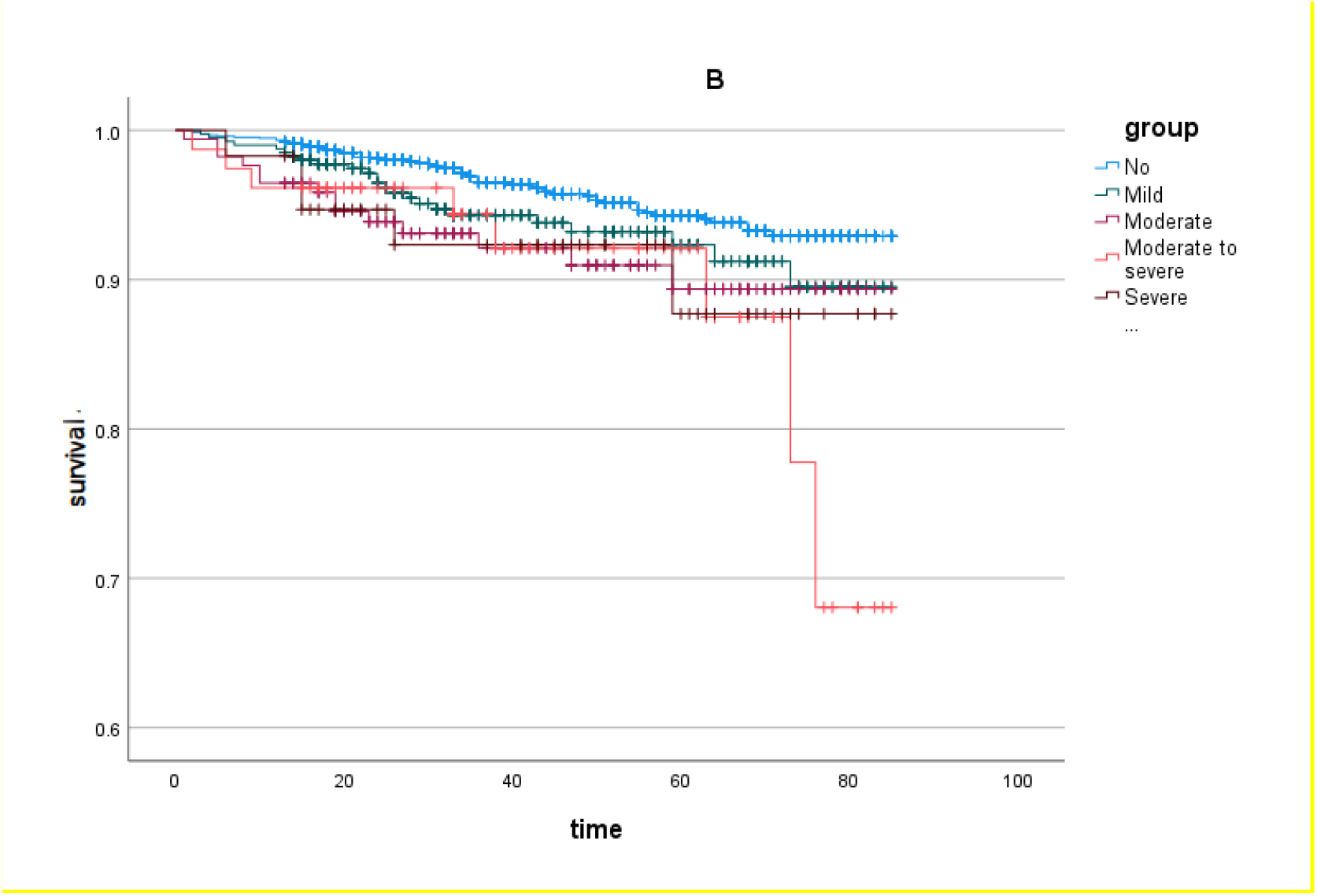
Kaplan-Meier survival curve of prediabetic patients with/without depression A. Kaplan-Meier survival curve of the prediabetes patients with/without depression; B. Kaplan-Meier survival curve of the prediabetes patients with different statuses of depression

## Discussion

Depression is a common mental illness. Its main characteristics include low mood, reduced interest, pessimism, slow thinking, lack of initiative, self-blame, poor diet, and sleep, worry about suffering from various diseases, and feeling that the whole body is tired. Feeling unwell, and in severe cases, suicidal thoughts and behaviors may occur. Maina et al. ^[5]^reported that there is a causal relationship and a positive correlation between depression and type 2 diabetes. Prediabetes is a state in which blood sugar levels have increased but have not yet reached the diagnostic criteria for diabetes. During this stage, an individual’s risk of developing diabetes increases. To explore this relationship further, our study aimed to examine the relationship between depression and prediabetes.

The results of this study indicate that moderate to severe depression is significantly associated with the development of prediabetes. Similarly, there may be a bidirectional relationship between prediabetes and anxiety and depression, that is, when a patient is diagnosed with prediabetes, it can aggravate the occurrence of anxiety and depression, and anxiety and depression can accelerate the risk of prediabetes converting into diabetes. ^[6]^ Zhou, Lu ^[7-9],^ and others reported that the incidence of anxiety and depression in patients with prediabetes is higher than that in the normal population. As early as 2016, Liu et al.^[10]^ established a one-year China Health and Retirement Longitudinal Study (CHARLS), which included Chinese elderly with known or unknown diabetes. Participants with newly diagnosed diabetes or prediabetes were considered to have unknown diabetes. Findings showed that knowledge of having diabetes, treatment, and other chronic conditions was associated with higher rates of depression in patients with known diabetes compared with those with unknown diabetes. The prevention of depression in patients with diabetes should receive more attention among middle-aged, female, and less educated people. Some of the findings are similar to ours in that we used the PHQ-9 criterion to stratify depressive status, which significantly contributes to the development of depression. Therefore, large prospective studies are still needed to study the impact of depression on the development and prognosis of prediabetes.

Multiple previous studies have shown ^[11-13]^ that major depression is significantly associated with mortality in type 2 diabetes. In our study, moderate to severe depression was found to be positively associated with all-cause mortality in patients with prediabetes. Increased hypothalamic-pituitary-adrenal (HPA) axis activity has been implicated in the pathophysiology of depression and anxiety, leading to the development of some metabolic precursors of type 2 diabetes (e.g., insulin resistance, hypertension, and visceral obesity) through increased cortisol exposure. Likewise, inflammatory markers and activation of the immune system are also thought to be links between depression and the development of diabetes. Depression is associated with the activation of several relevant biological systems, including the HPA axis, the sympathetic nervous system, and the inflammatory response, which can influence insulin resistance and glycemic outcomes. Further confounding the relationship between depression, anxiety, and the transition from prediabetes to type 2 diabetes. Medications used to treat these mental health disorders are associated with an increased risk of type 2 diabetes. Early screening and early intervention for patients taking these medications can significantly reduce the risk of prediabetes. Patient mortality. ^[14, 15]^

Recently, a genetic study used genetic correlation analysis, multigene overlap analysis, and Mendelian randomization analysis to confirm the intrinsic link between type 2 diabetes and attention deficit hyperactivity disorder. ^[16]^In the study, genetic correlation analysis showed that type 2 diabetes and attention deficit hyperactivity disorder showed a significant positive correlation in two gene sets (ADHD2019 and ADHD2022). Multigene overlap analysis showed that in the ADHD2022 gene set, type 2 diabetes and attention deficit hyperactivity disorder share the vast majority (90%) of risk variant genes, and Mendelian randomization analysis showed that in the ADHD2019 gene set, attention deficit hyperactivity disorder There is a genetic bidirectional causal relationship between ADHD and type 2 diabetes. The research results of Wang^[17]^ also proved that adults with a history of attention deficit hyperactivity disorder are strongly related to the occurrence of depression and alcoholism. Therefore, depression and attention deficit symptoms may be related to biology, metabolism, and life. It is related to many risk factors such as lifestyle, which lead to the progression of prediabetes to type 2 diabetes.

### Clinical significance

Early diagnosis and intervention of prediabetes are of great significance in preventing diabetic transformation and complications. Currently, the first-line treatment for prediabetes is lifestyle modification or metformin. Intensive lifestyle modifications such as diet, exercise, and weight loss are more beneficial than metformin use. However, there are currently no effective ways to reduce the incidence of prediabetes. We provide further valuable insights into elucidating the relationship between depression and prediabetes. Due to the fast pace of modern work and the high pressure in people’s lives, depression has become a relatively common mental illness. Our research report provides a new insight into the risk and treatment of prediabetes from the perspective of mental health conditions. Endocrinologists should focus on the mental health of patients with prediabetes, not just those with diabetes. Endocrinologists should not only pay attention to patients’ mental health status during diabetes treatment but also pay attention to preventing the development and worsening of prediabetes by paying attention to mental health status. Future research is warranted to investigate whether therapeutic interventions aimed at addressing mental health can reverse or impede the progression from prediabetes to diabetes, thereby preventing subsequent negative health outcomes.

## Conclusion

In conclusion, moderate to severe and severe depression was significantly associated with the risk of prediabetes. Moderate to severe depression is positively associated with mortality in patients with prediabetes. This suggests that depression may be a new and valuable indicator of prediabetes risk. Early treatment of depression improves outcomes in prediabetes.

## Data Availability

All data are available from NHANES database (survey Data and Documentation NANES 2013-2014,NHANES 2015-2016,NHANES 2017-2018.)

## Acknowledgments

The authors thank all the staff and participants in NHANES 2013–2018 for their contribution to the donation, collection, and sharing of data.

## Author contributions

JZ analyzed the data and drafted the manuscript. XY drafted the manuscript. All authors made substantial contributions to the interpretation of the data and provided critical revisions to the manuscript.

## Availability of data and materials

All data are available from NHANES database (survey Data and Documentation NHANES 2013-2014,NHANES 2015-2016,NHANES 2017-2018.).

## Declarations

### Ethics approval and consent to participate

This study was conducted by the Helsinki Declaration of 1975, as revised in 2013. NHANES 2013-2018 were approved by The NCHS Ethics Review Board, and informed consent was obtained from all participants. This study was based on publicly available data. The acquisition and analysis of data were consistent with NHANES research requirements.

